# Cellular immunity predominates over humoral immunity after the first dose of COVID-19 vaccines in solid organ transplant recipients

**DOI:** 10.1101/2021.05.07.21256809

**Authors:** Tina Schmidt, Verena Klemis, David Schub, Sophie Schneitler, Matthias C. Reichert, Heinrike Wilkens, Urban Sester, Martina Sester, Janine Mihm

## Abstract

Knowledge on the vaccine-induced cellular and humoral immunity and on immunogenicity of vector-based and mRNA vaccines in solid organ transplant recipients is limited. Therefore, SARS-CoV-2 specific T-cells and antibodies were analyzed in 40 transplant recipients and 70 age-matched controls after the first dose of vector-based or mRNA vaccines. Plasmablasts and SARS-CoV-2 specific CD4 and CD8 T-cells were quantified using flow-cytometry. Specific antibodies were analyzed by ELISA and neutralization assay. SARS-CoV-2 specific antibodies and T-cells were induced in both groups with significantly lower levels in patients. While antibodies were detected in 80% of controls and 5.3% of patients, specific CD4 and/or CD8 T-cells were more frequently found in both controls (84.3%) and patients (23.7%). The two vaccine types showed notable differences, as IgG and neutralizing activity were more pronounced after mRNA vaccination (p<0.0001 each), whereas CD4 and CD8 T-cell levels were higher after vector vaccination (p=0.009; p<0.0001). Plasmablast numbers were significantly higher in controls and correlated with SARS-CoV-2 specific IgG- and CD4 T-cell levels. In conclusion, assessment of antibodies is not sufficient to identify COVID-19-vaccine responders. Together with differences in immunogenicity among vaccines, this necessitates combined analysis of humoral and cellular immunity to reliably assess responders among immunocompetent and immunocompromised individuals.

## Introduction

Currently licensed vaccines towards the severe acute respiratory syndrome coronavirus type 2 (SARS-CoV-2) include mRNA vaccines and adenovirus-based replication-incompetent vector vaccines. Both vaccine types have proved to be strongly immunogenic and highly efficient in preventing severe coronavirus disease (COVID-19) in immunocompetent individuals (1-3). Immunocompromised individuals such as patients after solid organ transplantation are at higher risk to suffer from more severe courses of disease (4). Therefore, although this group of patients was not included in vaccine trials, COVID-19 vaccination is generally recommended for transplant recipients with no preference towards the use of either mRNA or vector-based vaccines (5). It is well known that humoral immunity towards other vaccines such as influenza or hepatitis B is decreased in solid organ transplant recipients (6-8). Likewise, early reports in transplant recipients immunized with mRNA-based COVID-19 vaccines showed that SARS-CoV-2-specific antibodies were only induced in approximately 6-17% after the first dose (9-11), and up to 59% after the second dose (12-17), respectively. Risk factors for poor response included older age, more intense immunosuppressive drug regimens including depleting antibodies and anti-metabolites, and earlier time after transplantation (9, 12). While the ability to induce antibodies has primarily been reported so far, vaccine-induced cellular immunity in transplant recipients is poorly studied. Moreover, immunogenicity in transplant recipients after administration of vector-based vaccines is currently unknown, and general knowledge on potential differences of immunogenicity between the two vaccine types is scarce. Although both the mRNA vaccines and the currently licensed vector vaccine ChAdOx1 nCoV-19 are administered twice, the recommended time interval between the first and the second dose varies from 3-6 weeks for mRNA vaccines to 9-12 weeks for the ChAdOx1 nCoV-19 vaccine (5).

Here we report the results of a prospective study assessing the vaccine-induced humoral and cellular immune response in solid organ transplant recipients in comparison with a healthy age-matched control group. The vector vaccine ChAdOx1 nCoV-19 and the mRNA vaccines BNT162b2 or mRNA-1273 were assigned as per national policies (5). To allow direct comparison of immunogenicity of the two vaccine types independent of the recommended time interval between the first and the second dose, the focus of this report addressed primary induction of humoral and cellular immunity after the first vaccine dose.

## Methods

### Study design and patient population

Solid organ transplant recipients and age-matched immunocompetent controls with no known history of SARS-CoV-2 infection were included in the study. Individuals either received one of the mRNA-vaccines (BNT162b2 or mRNA-1273) or the adenovirus-vector vaccine ChAdOx1 nCoV-19 as per national recommendation. Lymphocyte subpopulations as well as vaccine-induced SARS-CoV-2-specific humoral and cellular immune responses were analyzed from heparinized whole blood 13-30 days after the first vaccine dose. Analyses of lymphocyte subpopulations and antigen-specific T cells were carried out within 24 hours. Antibody testing was performed from frozen plasma samples. Baseline levels of SARS-CoV-2-reactive antibodies were determined to control for pre-existing immunity. Local and systemic adverse events within 7 days after vaccination were recorded using a questionnaire. The study was approved by the ethics committee of the Ärztekammer des Saarlandes (reference 76/20), and all individuals gave written informed consent.

### Quantification of lymphocyte populations and plasmablasts

Quantification of T cells, B cells and plasmablasts was performed on 100 µl heparinized whole blood as described before (18) using monoclonal antibodies towards CD3 (clone SK7), CD19 (clone HIB19), CD27 (clone L128), CD38 (clone HB7) and IgD (clone IA6-2). Prior to staining whole blood was washed with medium (RPMI) to remove soluble CD27 and IgD. After 25 min of incubation, samples were treated with lysing solution (BD Biosciences). Thereafter, cells were washed and analyzed using flow cytometry (FACS-Canto-II) and FACS-Diva-V6.1.3-software (BD Biosciences). Among total lymphocytes, T and B cells were identified by expression of CD3 and CD19, respectively. Plasmablasts were identified as CD38 positive cells among IgD-CD27+ CD19 positive switched-memory B cells. Differential blood counts were used to calculate absolute lymphocyte numbers.

### Quantification of vaccine induced SARS-CoV-2-specific T cells

SARS-CoV-2-specific T cells were determined as described previously (18). In brief, heparinized whole blood samples were stimulated with overlapping peptides spanning the SARS-CoV-2 spike protein (spike vial 1, N-terminal receptor binding domain and spike vial 2, C-terminal portion including the transmembrane domain, each peptide 2µg/ml; JPT, Berlin, Germany). Stimulations with the peptide diluent (0.64% DMSO) and with 2.5μg/ml of the polyclonal stimulus *Staphylococcus aureus* enterotoxin B (SEB; Sigma) served as negative and positive controls, respectively. All stimulations were carried out in presence of co-stimulatory antibodies against CD28 and CD49d (1μg/ml each). Cells were stimulated for 6 hours, processed as described before (18) and immunostained using anti-CD4 (clone SK3), anti-CD8 (clone SK1), anti-CD69 (clone L78), anti-IFNγ (clone 4S.B3), anti-IL-2 (clone MQ1-17H12), and anti-TNFα (clone MAb11). All markers were analyzed on a single-cell level using flow-cytometry. SARS-CoV-2-reactive CD4 or CD8 T cells were identified as activated CD69 positive T cells producing IFNγ. In addition, co-expression of IL-2 and TNFα was analyzed to characterize functionality in more detail. Reactive CD4 and CD8 T-cell levels after control stimulations were subtracted from levels obtained after SARS-CoV-2-specific stimulation, and 0.03% of reactive T cells was set as detection limit based on the distribution of T-cell frequencies after control stimulations.

### Determination of SARS-CoV-2-specific antibodies and neutralization capacity

SARS-CoV-2-specific IgG antibodies towards the receptor binding domain of SARS-CoV-2 spike protein were quantified using an enzyme-linked immunosorbent assay (ELISA) according to the manufacturer’s instructions (SARS-CoV-2-QuantiVac, Euroimmun, Lübeck, Germany). Antibody binding units (BAU/ml) <25.6 were scored negative, levels between ≥25.6 and <35.2 were scored intermediate, and levels ≥35.2 were scored positive. A neutralization assay based on antibody-mediated inhibition of soluble ACE2 binding to the plate-bound S1 receptor binding domain was used according to the manufacturer’s instructions (SARS-CoV-2-NeutraLISA, Euroimmun, Lübeck, Germany). Neutralizing antibodies were determined in all individuals with SARS-CoV-2 positive IgG titers. Neutralizing capacity is expressed as percentage of inhibition (IH) which is calculated by 1 minus the ratio of the extinction of the patient sample and the extinction of the blank value. IH<20% were scored negative, IH≥20 to <35 were scored intermediate, and IH≥35% were scored positive.

### Statistical analysis

Mann-Whitney test was performed to compare unpaired non-parametric data between groups (time since vaccination, lymphocyte subpopulations, T-cell and antibody levels). Data with normal distribution were analyzed using unpaired t test (cytokine-expression profiles, age). Categorial analyses on gender, type of vaccine, T-cell responses, and adverse events were performed using Fisher’s exact test. Correlations between levels of T cells, antibodies, and plasmablasts were analyzed according to Spearman. A p-value <0.05 was considered statistically significant. Statistical analysis was carried out using GraphPad Prism 9.0 software (GraphPad, San Diego, CA, USA) using two-tailed tests.

## Results

### Study population

A total of 40 solid organ transplant recipients and 70 immunocompetent controls were recruited and tested at a median of 15 (IQR 6) days after the first vaccination with either vector vaccine or mRNA vaccine, which was applied in 75% and 25% of individuals, respectively. All individuals had no known history of SARS-CoV-2 infection. The two groups were matched regarding age, and vaccine type, but there were more males among patients (table 1). Patients were mainly kidney recipients transplanted for 6.5 (IQR 9.9) years, and the majority of patients (28/40, 70%) were on an immunosuppressive triple drug regimen including a calcineurin inhibitor, glucocorticoids and an antimetabolite (table 1). Patients had a leukocytosis mainly affecting neutrophils and had significantly less lymphocytes. Among lymphocytes, a trend towards lower numbers of CD3 T cells, and significantly lower numbers of CD19 positive B-cell numbers were observed in patients (table 1).

**Table 1:** Demographic and clinical characteristics of the study population.

|  | Controls | transplant recipients |  |
| --- | --- | --- | --- |
|  | n=70 | n=40 | p-value |
| **Years of age (mean±SD)** | 50.6±11.9 | 54.5±12.7 | 0.11 |
| **Female gender, n (%)** | 49 (70.0) | 18 (45.0) | 0.01 |
| **Vaccine (vector vs. mRNA)** |  |  | 0.654 |
| vector (ChAdOx1 nCoV-19), n (%) | 51 (72.9) | 31 (77.5) |  |
| mRNA, n (%) | 19 (27.1) | 9 (22.5) |  |
| *BNT162b2*, n (%) | 14 (20.0) | 8 (20.0) |  |
| *mRNA-1273*, n (%) | 5 (7.1) | 1 (2.5) |  |
| Analysis time[days after vaccination], median (IQR) | 15 (5) | 16 (7) | 0.166 |
| **Organ transplant** |  |  |  |
| Kidney, n (%) | n.a. | 36 (90.0) | n.a. |
| Heart, n (%) | n.a. | 1 (2.5) | n.a. |
| Liver, n (%) | n.a. | 1 (2.5) | n.a. |
| Liver and kidney, n (%) | n.a. | 2(5.0) | n.a. |
| Years since transplantation, median (IQR) | n.a. | 6.5 (9.9) | n.a. |
| **Immunosuppressive regimen** |  |  |  |
| CNI, antimetabolite, GC (%) | n.a. | 28 (70.0) | n.a. |
| CNI, antimetabolite (%) | n.a. | 5 (12.5) | n.a. |
| CNI, GC (%) | n.a. | 2 (5.0) | n.a. |
| CNI (%) | n.a. | 2 (5.0) | n.a. |
| mTOR-I, antimetabolite, GC (%) | n.a. | 2 (5.0) | n.a. |
| mTOR-I, GC (%) | n.a. | 1 (2.5) | n.a. |
| **Differential blood cell counts** |  |  |  |
|  | n=68 | n=38 |  |
| Leukocytes (cells/μl), median (IQR) | 6450 (1975) | 7600 (3275) | 0.017 |
| Granulocytes (cells/μl), median (IQR) | 3716 (1548) | 4710 (2519) | 0.001 |
| Monocytes (cells/μl), median (IQR) | 557 (212) | 627 (457) | 0.089 |
| Lymphocytes (cells/μl), median (IQR) | 2126 (831) | 1526 (1093) | 0.004 |
| CD3 T cells (cells/μl), median (IQR)* | 1471 (632) | 1101 (1081) | 0.061 |
| CD19 B cells (cells/μl), median (IQR)* | 201 (139) | 72 (124) | <0.0001 |

CNI: calcineurin inhibitor; antimetabolite: azathioprine or mycophenolate mofetil/mycophenolic acid; GC: glucocorticoids; mTOR-I: m-TOR inhibitor; \*B and T-cell counts were calculated on 56 controls and 37 transplant recipients.

### Lower SARS-CoV-2-specific antibody and T-cell levels in transplant recipients

When evaluating antibody responses after vaccination, spike-specific IgG levels were induced in 56 out of 70 immunocompetent controls (80.0%). In contrast, antibody titers were significantly lower in transplant recipients with most patients remaining seronegative (figure 1A, p<0.0001). Of note, the two kidney transplant recipients with the highest antibody titers (6511 and 292 BAU/ml, respectively) already had detectable SARS-CoV-2 antibodies at baseline (384 and 306 BAU/ml, respectively), which identified a history of an asymptomatic SARS-CoV-2 infection. The results of these patients are displayed separately from the remaining baseline-negative transplant recipients, of which only 2 patients each had vaccine-induced positive and intermediate titers, respectively (figure 1A). As expected from IgG titers, neutralizing antibody activity was only detected among healthy controls, of which only 18/70 (25.7%) reached an inhibitory capacity ≥35% (figure 1A). Among transplant recipients, neutralizing antibodies were only found in the two patients with prior infection, who had strong neutralizing activity (100% and 72.9% inhibition, respectively).

**Figure 1:**
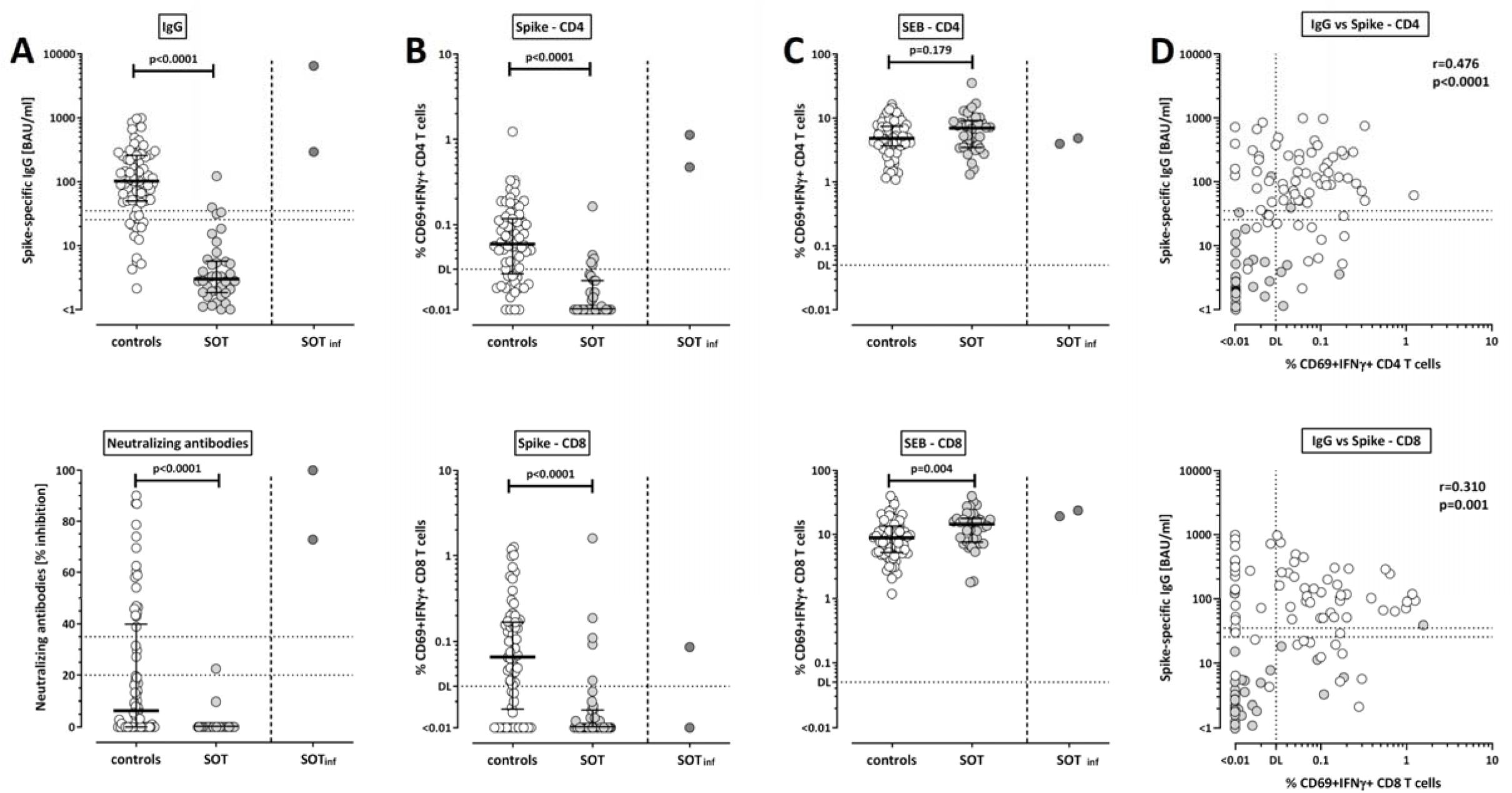
T-cell and antibody responses in controls and transplant recipients after SARS-CoV-2-specific vaccination. **(A)** Levels of spike-specific IgG and neutralizing antibodies were determined by ELISA and neutralization assay, respectively, and compared between healthy controls (open symbols) and solid organ transplant recipients (SOT, filled symbols). Levels of SARS-CoV-2-specific **(B)** and SEB-reactive **(C)** CD4 and CD8 T cells were flow-cytometrically determined after antigen-specific stimulation in vitro followed by intracellular cytokine staining, and compared between groups. In panels A-C, results of two vaccinated transplant recipients with a previously unknown history of asymptomatic SARS-CoV-2 infection are displayed separate from the remaining baseline-negative transplant recipients and excluded from statistical analyses. Bars represent medians with interquartile ranges. Differences between the groups were calculated using Mann-Whitney test. **(D)** Correlations between IgG titers and SARS-CoV-2-specific CD4 and CD8 T cells are shown (calculated according to Spearman). Dotted lines indicate limits for IgG and neutralizing antibodies, including negative, intermediate and positive results, respectively as per manufacturer’s instructions, or detection limits (DL) for SARS-CoV-2-specific and SEB-reactive CD4 and CD8 T cells, respectively. IFN, interferon.

To evaluate cellular immune responses after vaccination, heparinized whole blood samples were stimulated with overlapping peptide pools of the SARS-CoV-2 spike protein followed by intracellular staining of IFNγ. Polyclonal T-cell reactivity in SEB-stimulated samples served as positive controls. SARS-CoV-2- and SEB-reactive CD4 and CD8 T cells were identified by co-expression of the activation marker CD69 and IFNγ (figure 1B and C). In line with antibody levels, both vaccine-induced SARS-CoV-2-specific CD4 and CD8 T-cell levels were significantly higher in healthy controls compared with solid organ transplant recipients (figure 1B, each p<0.0001). This was not due to a general non-responsiveness, as CD4 and CD8 T-cell reactivity after polyclonal stimulation was not impaired in transplant recipients (figure 1C). Overall, SARS-CoV-2-specific IgG levels showed a significant correlation with specific CD4 (r=0.476, p<0.0001) and CD8 T cells (r=0.310, p=0.001, figure 1D). The two transplant recipients with positive SARS-CoV-2 antibody titers at baseline showed a pronounced SARS-CoV-2-specific cellular immunity, which was similar or even higher in magnitude as compared to vaccine responses in immunocompetent naïve individuals.

The percentages of individuals with cellular and humoral immunity above the respective detection limits is summarized in table 2. Fifty-six out of 70 controls (80.0%) and only 2/38 (5.3%) of transplant recipients had IgG levels above threshold (table 2). In contrast, a larger fraction of patients showed a vaccine-induced SARS-CoV-2-specific cellular immune response (9/38 (23.7%) transplant recipients and 59/70 (84.3%) controls). Finally, 67/70 (95.7%) controls and 10/38 (26.3%) of transplant recipients mounted any type of vaccine-induced immune response (antibodies and/or T cells).

**Table 2:** Vaccine-induced immune responses and reported adverse events in the study population.

|  | Controls<br>n=70 | transplant recipients<br>n=38* | p-value |
| --- | --- | --- | --- |
| **Individuals with SARS-CoV-2 spike-specific immunity** |  |  |  |
| IgG ($\geq 35.2$ BAU/ml), n (%) | 56 (80.0) | 2 (5.3) | <0.0001 |
| neutralizing antibodies (IH$\geq 35\%$), n (%) | 18 (25.7) | 0 (0) | 0.0002 |
| CD4 T cells, n (%) | 52 (74.3) | 5 (13.2) | <0.0001 |
| CD8 T cells, n (%) | 49 (70.0) | 5 (13.2) | <0.0001 |
| **Combined cellular and/or humoral analysis** |  |  |  |
| CD4 and/or CD8 T cells, n (%) | 59 (84.3) | 9 (23.7) | <0.0001 |
| T cells and/or IgG; n (%) | 67 (95.7) | 10 (26.3) | <0.0001 |
| **Adverse events** |  |  | 0.018<sup>#</sup> |
| none, n (%) | 8 (11.4) | 12 (31.6) |  |
| local, n (%) | 11 (15.7) | 4 (10.5) |  |
| systemic, n (%) | 17 (24.3) | 11 (28.9) |  |
| local and systemic, n (%) | 34 (48.6) | 11 (28.9) |  |

Ab, antibody; BAU: binding antibody units; IH: percentage of inhibition; \*the two patients with evidence of prior infection are not included in this table; <sup>#</sup>comparison of none versus any adverse event (Fisher's exact test), for differences in adverse events between the vaccines see figure 4.

### Differential induction of SARS-CoV-2-specific T cells and antibodies after vector-based vaccine and mRNA vaccines

As two types of vaccines were administered in our study, antibody and T-cell responses in controls and patients were stratified according to vector and mRNA vaccines. As shown in figure 2A, induction of SARS-CoV-2-specific IgG and neutralizing antibodies in healthy controls was far more pronounced after immunization with the mRNA vaccines (median IgG 264.1 (IQR 302.2) BAU/ml) as compared to titers after vector-based vaccination (median IgG 75.3 (IQR 112.7) BAU/ml, p<0.0001). This contrasts with cellular immunity, where both SARS-CoV-2-specific CD4 and CD8 T-cell levels were significantly higher after vector vaccination (0.072% of CD4 T cells and 0.106% of CD8 T cells) as compared to mRNA vaccination (0.021% of CD4 T cells, p=0.009, and 0.007% of CD8 T cells, p<0.0001, figure 2B). In transplant recipients, neutralizing titers of very low inhibitory activity were found exclusively in two mRNA vaccinated patients and SARS-CoV-2-specific CD4 T cells only occurred in five patients after vector vaccination. Apart from this, there were no striking differences between the vaccines in transplant recipients, which may result from a lower general immune reactivity in patients. Likewise, no differences were observed for SEB-reactive T cells (figure 2C).

**Figure 2:**
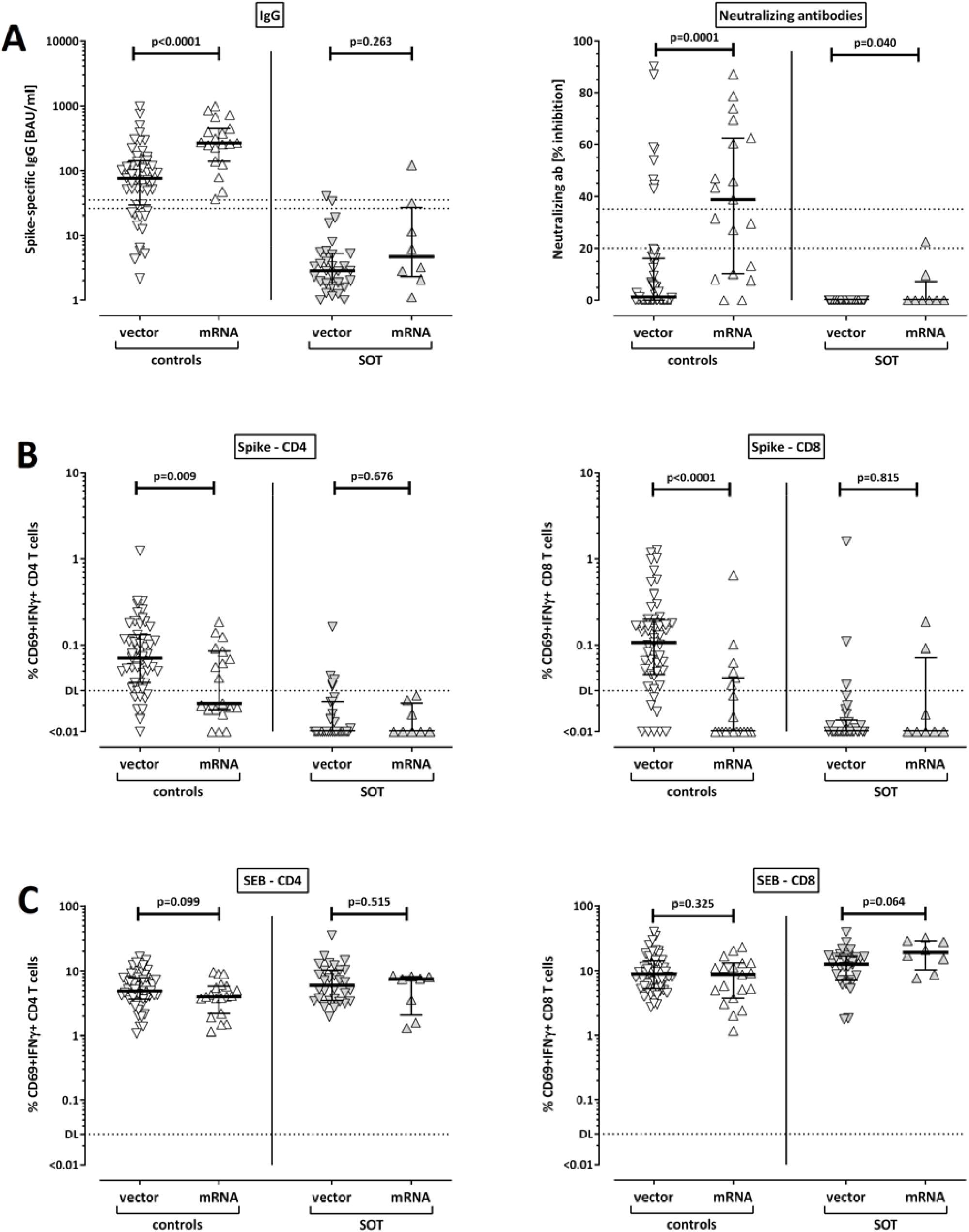
SARS-CoV-2-specific antibodies and T cells stratified according to vector and mRNA vaccines. **(A)** Levels of SARS-CoV-2-specific IgG and neutralizing antibodies, as well as levels of SARS-CoV-2-specific **(B)** and SEB-reactive **(C)** CD4 and CD8 T cells were compared among controls (open symbols) or among solid organ transplant recipients (SOT, filled symbols). Bars represent medians with interquartile ranges. Differences between the groups were calculated using Mann-Whitney test. Dotted lines in (A) indicate limits for IgG and neutralizing antibodies, including negative, intermediate and positive results, respectively as per manufacturer’s instructions. In (B) and (C) dotted lines indicate detection limits (DL) for SARS-CoV-2-specific and SEB-reactive CD4 and CD8 T cells. IFN, interferon.

Given the quantitative differences in cellular immune responses in individuals after mRNA and vector vaccination, we also analyzed qualitative differences in specific T-cell responses based on cytokine profiling of IFNγ, IL-2 and TNFα in individuals with detectable specific immunity. This analysis was restricted to individuals who had at least 30 cytokine positive events which included 46 controls (34 after vector and 12 after mRNA vaccination, respectively). As shown in figure S1, most cells after vector vaccination were multifunctional simultaneously producing all three cytokines, followed by cells producing IL-2 and TNFα. After mRNA vaccination, the percentage of triple positive CD4 T cells was significantly lower, and the percentage of IL-2/TNFα producing cells was significantly higher compared to cells after vector vaccination. CD8 T cells were dominated by IFNγ and TNFα production and showed only marginal IL-2 production (data not shown). As specific CD8 T cells were predominantly found among individuals after vector vaccination (figure 2B), this precluded a meaningful comparison of CD8 T-cell cytokine profiles with mRNA responses.

### The number of plasmablasts correlates with SARS-CoV-2-specific antibody and T-cell levels

We have previously shown that induction of SARS-CoV-2-specific immunity after natural infection was associated with an expansion of plasmablasts (18). To elucidate whether this is also observed after vaccination, vaccine-induced plasmablasts were quantified from whole blood as CD38 positive cells among IgD-CD27+ CD19 positive switched-memory B cells. As shown from 15 healthy controls analysed before and after vaccination, there was a significant increase from 0.88/µl before vaccination to 1.29/µl after vaccination (p=0.022, data not shown). In the whole group, controls had significantly higher numbers of vaccine-induced plasmablasts compared to transplant recipients (p<0.0001, figure 3A). In line with higher IgG titers in mRNA-vaccinated controls, median numbers of plasmablasts after mRNA vaccination were also higher than after vector vaccination, although the difference did not reach statistical significance (figure 3B). Plasmablast numbers also showed a correlation with vaccine-induced IgG titers (r=0.459, p<0.0001) as well as with SARS-CoV-2-specific CD4 T cells (r=0.358, p=0.0005, figure 3C). As expected, no correlation was found with specific CD8 T cells (p=0.265, data not shown).

**Figure 3:**
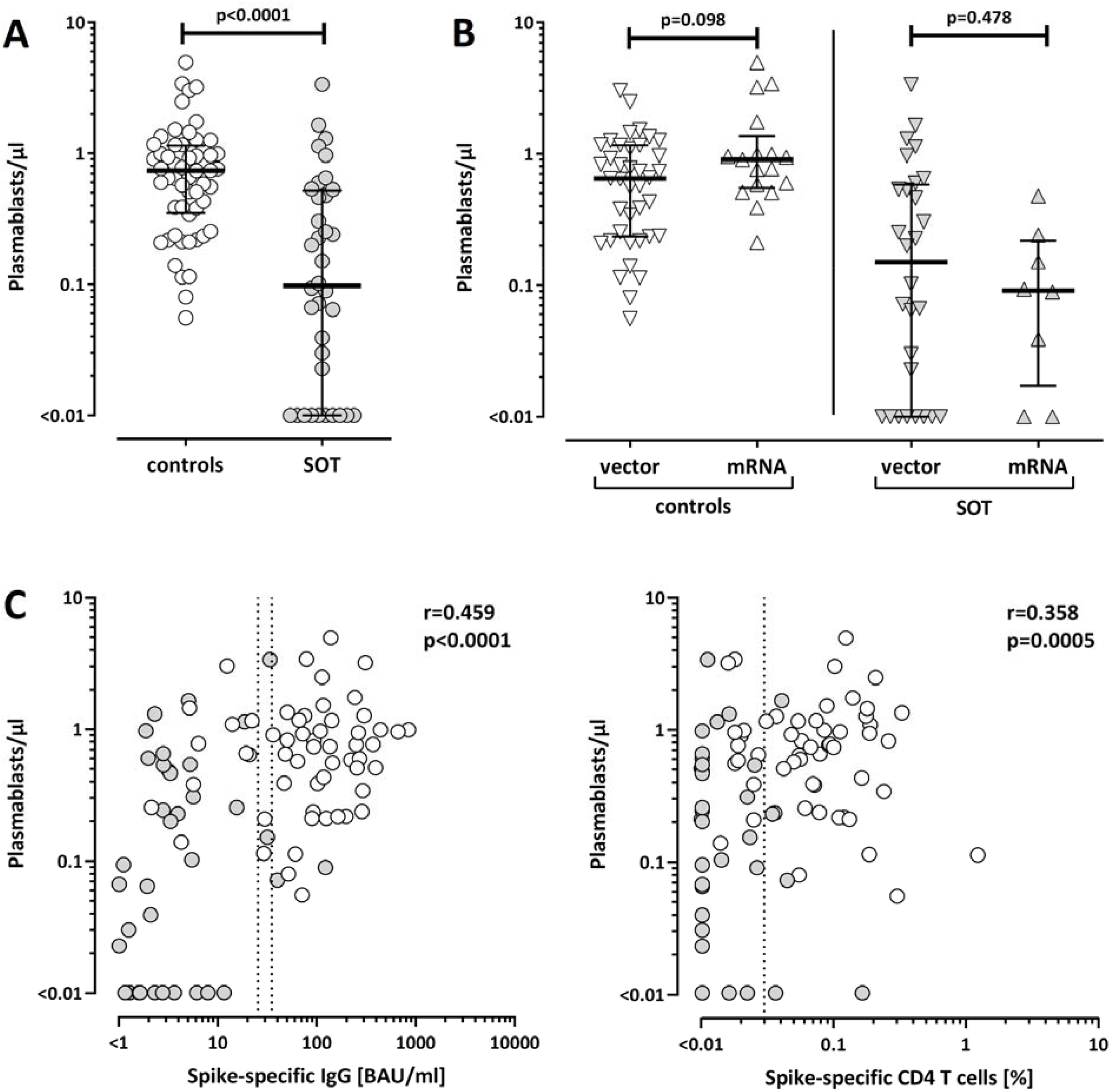
Levels of plasmablasts in healthy controls and transplant recipients after SARS-CoV-2 vaccination and correlation with antibody and T-cell responses. Numbers of plasmablasts were compared **(A)** between healthy controls (n=56) and solid organ transplant recipients (SOT, n=36) and **(B)** between individuals after immunization with vector-based or mRNA vaccines. **(C)** Plasmablasts were correlated with levels of SARS-CoV-2 spike-specific IgG and CD4 T cells. Bars in (A) and (B) represent medians with interquartile ranges. Differences between the groups were calculated using Mann-Whitney test. Correlations in (C) were analyzed according to Spearman. Dotted lines indicate (left panel) detection limits for IgG, indicating negative, intermediate and positive levels, respectively as per manufacturer’s instructions or (right panel) detection limit for SARS-CoV-2-specific CD4 T cells.

### Transplant recipients reported less adverse events after vaccination than healthy controls

Adverse events in the first week after vaccination were recorded using a questionnaire. As shown in table 2, the vaccine was better tolerated among transplant recipients; 32% of them reported neither local nor systemic symptoms, whereas this was the case in only 11% of controls (p=0.018). There was no difference in the occurrence of systemic adverse events between the two groups. However, the vector vaccine caused more systemic adverse events than the mRNA vaccine, which only reached statistical significance in controls (p=0.033, figure 4A). It is interesting to note, that systemic adverse events may be linked to cellular immunity as individuals with systemic symptoms had significantly higher levels of vaccine-induced SARS-CoV-2-specific CD8 T cells as compared to individuals who only had local or no symptoms (p=0.032, figure 4B).

**Figure 4:**
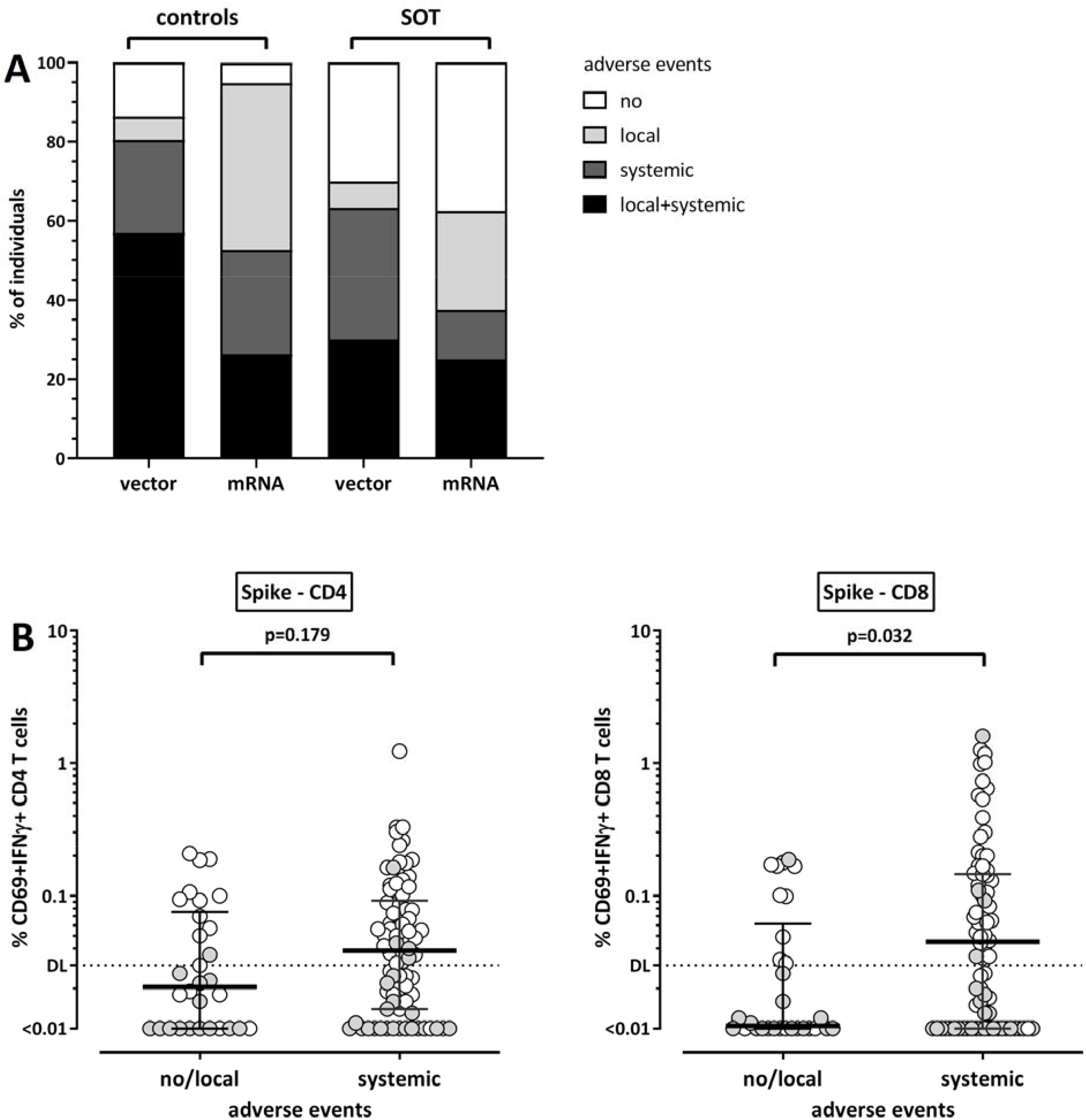
Increased levels of SARS-CoV-2-specific CD8 T cells in individuals with systemic adverse events after vaccination. **(A)** Controls and solid organ transplant recipients (SOT) were subdivided according to occurrence of adverse events (no, local, systemic, local and systemic) after immunization with vector-based or mRNA vaccines. Local adverse events included pain, swelling or redness at the injection site. Systemic adverse events included tiredness, fatigue, fever, headache, chills, myalgia, arthralgia, nausea, vomiting, diarrhea, dizziness, and or allergic reactions. Controls after vector vaccination reported more systemic adverse events than controls after mRNA vaccination (p=0.033, Fisher’s test). **(B)** SARS-CoV-2-specific CD4 and CD8 T-cell levels were compared between individuals with no or local adverse events and individuals with systemic symptoms. Controls and transplant recipients are denoted with white and grey symbols, respectively. Bars represent medians with interquartile ranges. Differences between the groups were calculated using Mann-Whitney test. Dotted lines indicate detection limits for SARS-CoV-2-specific T cells.

## Discussion

Based on the lack of efficacy trials, knowledge on the adaptive immune response after COVID-19 vaccination in transplant recipients will have potential to provide guidance for clinical practice. Up to now, data in this patient population are limited to humoral immunity after mRNA vaccination (9-17), and no data exist on cellular immunity or vaccine-induced immunity in patients after vector-based vaccination. In this study, we show that SARS-CoV-2-specific antibodies and/or T cells are detectable in 95.7% of immunocompetent controls and 26.3% of transplant recipients after a single dose of mRNA-or vector-based vaccine. Although the majority of controls had specific immunity well above the detection limit, antibody titers were higher after mRNA vaccination, whereas the vector vaccine elicited higher levels of T cells. As expected, both antibody and T-cell levels were significantly lower in transplant recipients. Our data show that assessment of humoral immune responses is not sufficient to identify vaccine responders, whereas combined analysis with cellular immunity was superior among both immunocompetent and immunocompromised individuals. Finally, we show that a single vaccine dose in transplant recipients with a history of asymptomatic SARS-CoV-2 infection led to a strong booster effect on T cells and antibodies with neutralizing function, which was similar or even higher in magnitude as primary induction of vaccine immunity in healthy controls.

Impaired immunogenicity in transplant recipients is well known for a variety of currently licensed vaccines such as hepatitis B or influenza (6-8). Early reports after the first dose of COVID-19 mRNA vaccines also show evidence for poor humoral immunity with detectable antibody responses in only 6-17% of patients (9-11), although no control group was studied in parallel. As age is a major factor that adversely affects immunogenicity after COVID-19 vaccination also in the general population (19), inclusion of age-matched controls is essential to evaluate immunogenicity in a real-world setting. Our study also revealed a poor antibody response transplant recipients predominantly immunized with the ChAdOx1 nCoV-19 vector vaccine, but it should be emphasized that lack of seroconversion was also observed in 20% of age-matched controls. Up to now, no data exist on the combined analysis of humoral and cellular immunity in transplant recipients after the first dose of mRNA or vector vaccine. Our results show that exclusive assessment of humoral immunity leads to a considerable underestimation of true vaccine responses. Among the 10 patients with detectable immunity, only 2 showed antibody titers above detection limit, while 9 patients were identified as CD4 and/or CD8 T-cell positive. Underestimation based on humoral immunity may be particularly pronounced in individuals after ChAdOx1 nCoV-19 vector vaccination, where antibody titers in healthy controls were even lower than after mRNA vaccination. Conversely, as T-cell levels were higher after vector vaccination, analysis of cellular immunity may be superior in identifying responders. This is illustrated by the fact that all transplant recipients with detectable CD4 T cells had been immunized with the ChAdOx1 nCoV-19 vaccine.

Our study revealed marked differences in quantitative and qualitative characteristics of cellular and humoral immunity induced by the two vaccine types. This difference in immunogenicity may result from the fact that the mRNA vaccines are delivered as lipid nanoparticles, whereas the adenovirus vector vaccine more closely mimics a viral infection. The adjuvant effect of the viral vector may be stronger as compared to immunomodulatory mRNA modifications predominantly triggering intracellular Toll-like receptors (20). In our study, vector vaccination was associated with more systemic adverse events, which was more frequent in individuals with strong CD8 T-cell responses. Thus, a vector-induced inflammatory response may favor an efficient priming and expansion of T cells with potentially different induction kinetics. In support of an accelerated expansion and functional maturation, T-cell levels after vector vaccination were not only significantly higher but also had a higher percentage of multifunctional T cells. Conversely, mRNA-induced T-cell levels were lower with lower fractions of multifunctional and higher fractions of dual-cytokine producing cells. On the B-cell/antibody axis, we observed significant differences in the number of circulating plasmablasts, which directly correlated with spike-specific IgG and CD4 T cells. Among controls, plasmablasts were numerically higher in mRNA vaccinated patients which may directly be linked to higher titers of IgG and neutralizing antibodies observed in our study. While longitudinal studies will be required to further elucidate the basic mechanisms underlying the different response patterns over time, this knowledge is important when evaluating vaccine-induced immunity in patient groups on different COVID-19 vaccine regimens. First reports on humoral immunity after the second dose of an mRNA vaccine have shown that antibody titers were still lower as in controls, but reached detection limits in 38-59% of transplant recipients (12-15). While this clearly exceeds response rates after the first dose, follow-up studies including both antibodies and T cells will be instrumental in comprehensively evaluating immunogenicity after a full vaccination cycle. Given the differences in the dominance of cellular and humoral immunity, it is tempting to speculate whether heterologous vaccine regimens may lead to a more comprehensive booster response exceeding that of a series of identical vaccines. As a heterologous regimen is now recommended in Germany for all ChAdOx1 nCoV-19 vaccinated individuals below the age of 60 years (21), this will offer a unique possibility for comparative analysis of immunogenicity in both patients and controls.

Safety and immunogenicity trials of the licensed vaccines have reported more frequent systemic adverse events in individuals immunized with the vector vaccine compared to mRNA vaccines, where adverse events predominated after the second vaccination (22, 23). In line with recent observations in transplant recipients after the first and second dose of the mRNA vaccine (24, 25), patients after mRNA vaccination only had mild to moderate symptoms that were largely restricted to local pain at the injection site. Knowledge on adverse events was now extended to transplant recipients after vector vaccination, who reported more systemic symptoms, although these were less frequent than in controls.

Our study is limited by a small sample size of patients that included two different types of vaccines after the first vaccine dose only. Consequently, this study does not inform on immune responsiveness and on the booster immune responses after completion of the recommended two-dose regimens. On the other hand, inclusion of both vaccine types in one study allowed direct comparison of primary immunogenicity in vaccine naïve individuals independent from variabilities resulting from vaccine-specific differences in the time interval between the first and the second dose. As a strength of our study, we comprehensively assessed both the humoral immune response with IgG and neutralization capacity, and the cellular immune response that included both CD4 and CD8 T cells and their cytokine profiles. Inclusion of an age-matched control group is considered as an additional asset to provide comparative data on immunogenicity of immunocompetent individuals in a real-world setting.

In conclusion, we show that serology considerably underestimates immunogenicity of COVID-19 vaccines in both immunocompetent controls and transplant recipients, with an improved ability to identify vaccine responders after combined analysis of humoral and cellular immunity. Despite this advantage, the proportion of responders after the first vaccine dose among transplant recipients is significantly lower than among controls, which may result in higher risk of post-vaccination infection (26) and emphasizes the need to advocate for continued adherence to hand hygiene, physical distancing, and use of facemasks. On the other hand, as with immunocompetent individuals (27, 28), one vaccine dose may be sufficient in transplant recipients with prior infection, as pre-existing SARS-CoV-2-specific immunity was boosted to levels comparable with healthy controls. Regarding secondary vaccination, transplant recipients may benefit from the closest interval possible to ensure a timely booster effect of specific immunity. In contrast, current practice to delay the second dose to the longest interval in immunocompetent individuals is reasonable in times of limited vaccine supply. Finally, despite efficacy of one-dose vaccine types in immunocompetent individuals (29), it seems reasonable to preferentially advocate for two-dose regimens in vulnerable patient groups such as transplant recipients.

## Data Availability

All figures have associated raw data. The data that support the findings of this study are available from the corresponding author upon reasonable request.

## Abbreviations

BAU: antibody binding units
COVID-19: coronavirus disease 2019
DL: detection limit
ELISA: enzyme-linked immunosorbent assay
IFN: interferon
IH: percentage of inhibition
IL: interleukin
SARS-CoV-2: Severe acute respiratory syndrome coronavirus type 2
SEB: *Staphylococcus aureus* enterotoxin B
TNF: tumor necrosis factor

## Author Contributions

D.S., T.S., U.S., S.S., and M.S. designed the study; D.S., T.S., U.S. and M.S. designed the experiments, V.K., and T.S. performed experiments; S.S., H.W., M.C.R., J.M., and U.S. contributed to study design, patient recruitment, and clinical data acquisition. D.S., V.K., T.S., U.S., J.M. and M.S. supervised all parts of the study, performed analyses and wrote the manuscript. All authors approved the final version of the manuscript.

## Acknowledgements

The authors thank Candida Guckelmus and Rebecca Urschel for excellent technical assistance, and Susanne Brehmer, Inna Vallar, Fabio Lizzi and the team of the Saarland University Transplant center for their valuable support. The authors also thank all participants to this study who contributed to the gain in knowledge from this project. Financial support was given by the State Chancellery of the Saarland.

## Supplementary Information

**Figure S1:**
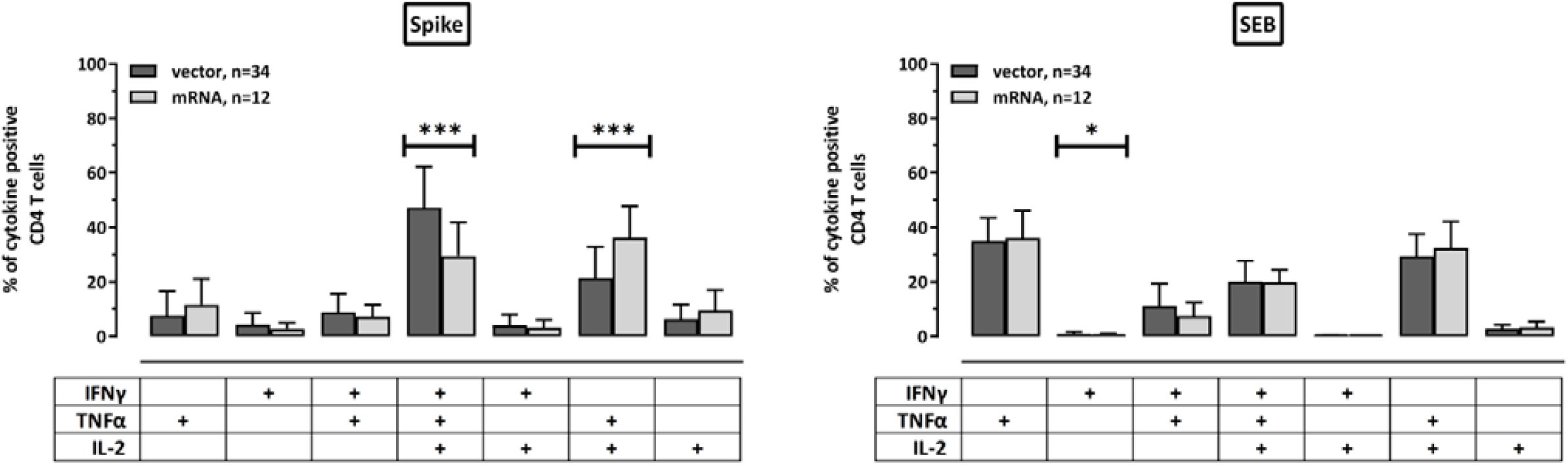
Different cytokine expression profiles of SARS-CoV-2-specific CD4 T cells after vector and mRNA vaccine. Cytokine expression of SARS-CoV-2-specific and SEB-reactive CD4 T cells was compared between healthy controls after immunization with vector or mRNA vaccines. Cytokine expressing CD4 T cells were divided into 7 subpopulations according to their expression of IFNγ, IL-2, and TNFα (single, double or triple cytokine expressing cells). To ensure robust statistical analysis in this setting, analysis was restricted to samples with at least 30 cytokine expressing CD4 T cells (n=34 for vector and n=12 for mRNA vaccines). Differences among subpopulations between controls with vector and mRNA vaccines were determined using unpaired t-test. Bars represent means and standard deviations of subpopulations among all SARS-CoV-2-specific or SEB-reactive CD4 T cells. IFN, interferon, IL, interleukin, TNF, tumor necrosis factor; *, p<0.05; ***, p<0.0001.

